# Association between the choroid plexus volume and cognitive function in community-dwelling older adults without dementia: A population-based cross-sectional analysis

**DOI:** 10.1101/2024.08.13.24311800

**Authors:** Yosuke Hidaka, Mamoru Hashimoto, Takashi Suehiro, Ryuji Fukuhara, Tomohisa Ishikawa, Naoko Tsunoda, Asuka Koyama, Kazuki Honda, Yusuke Miyagawa, Kazuhiro Yoshiura, Seiji Yuuki, Naoto Kajitani, Shuken Boku, Kazunari Ishii, Manabu Ikeda, Minoru Takebayashi

## Abstract

**Background:** Choroid plexus (CP) volume increase may be associated with cognitive decline in older individuals without dementia. In this study, we aimed to clarify whether CP volume can be an imaging marker of cognitive decline in older adults, verify its association with cognitive decline in older individuals compared with brain parenchymal and CSF volumes, and explore the factors associated with the CP volume.

**Methods:** We measured the CP volume, brain parenchyma, and cerebrospinal fluid (CSF) spaces associated with disproportionately enlarged subarachnoid space hydrocephalus (DESH), an imaging feature of normal-pressure hydrocephalus, in community-dwelling individuals aged ≥65 years without dementia.

**Results:** In 1,370 participants, lower Mini-Mental State Examination (MMSE) scores were significantly associated with higher CP volume, even after adjusting for DESH-related CSF space and brain parenchymal volume. CP volume was more strongly associated with MMSE scores than DESH-related CSF space and brain parenchymal volume. History of smoking, white matter hyperintensity, enlarged perivascular spaces, age, body mass index, and diabetes mellitus were associated with increased CP volume.

**Conclusions:** In community-dwelling older individuals without dementia, CP volume is associated with cognitive decline independent of the brain parenchyma and CSF volumes and may be one of the most sensitive imaging markers of cognitive decline. Our findings emphasize the importance of investigating CP volume increase to maintain cognitive function in older individuals. Further longitudinal studies are required.

**WHAT IS ALREADY KNOWN ON THIS TOPIC:** Choroid plexus (CP) volume increase has been associated with cognitive decline in older individuals without dementia; however, the influence of volumes of cerebrospinal fluid (CSF) and brain parenchyma in this association remains unexplored. The extent to which CP volume is involved in cognitive decline in older adults compared with brain parenchymal and CSF volumes is unclear.

**WHAT THIS STUDY ADDS:** CP volume is associated with cognitive decline in older individuals independent of the volumes of CSF and brain parenchyma. CP is one of the structures most closely related to cognitive impairment in older individuals without dementia.

**HOW THIS STUDY MIGHT AFFECT RESEARCH, PRACTICE OR POLICY:** Our study provides a reference for investigating CP volume increase to maintain cognitive function in older individuals.

## INTRODUCTION

The number of older individuals with dementia is increasing worldwide. It is important to identify older individuals with declining cognitive function at an early stage and guide them to preventive and therapeutic measures. However, preventive or disease-modifying therapies for cognitive impairment in older adults are currently limited, and deeper insights into the pathophysiology of cognitive impairment in older individuals are required. The relationship between intracranial structures other than brain parenchyma and cognitive decline in older adults has not been well studied.

In our previous study, [1] we performed volumetric quantification of cerebrospinal fluid (CSF) spaces associated with disproportionately enlarged subarachnoid space hydrocephalus (DESH) [2] using magnetic resonance imaging (MRI) data from community-dwelling older individuals. DESH is defined as tight high-convexity and medial subarachnoid spaces and enlarged Sylvian fissure (SF) with ventriculomegaly (Figure 1) and is a neuroimaging phenotype of idiopathic normal-pressure hydrocephalus (iNPH), which originates from impaired CSF dynamics. [2] We previously reported that the CSF space in DESH-related regions (e.g., ventricular system (VS), SF, and subarachnoid spaces at high convexity and midline [SHM]) (Figure 2) is an important structure associated with cognitive decline in older adults. [1]

**Figure 1.**
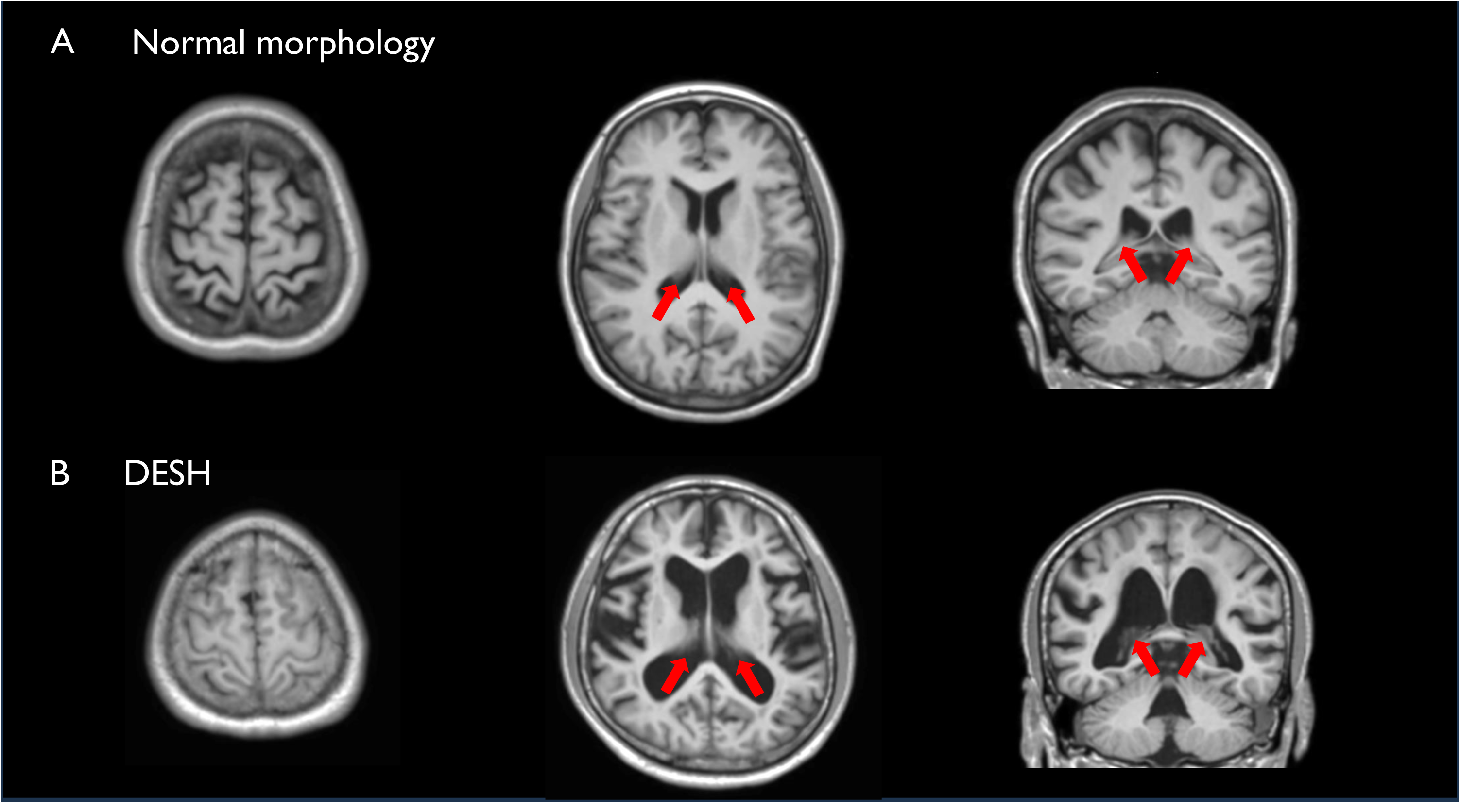
Comparison of brain magnetic resonance imaging (MRI) scans in a participant with normal morphology and a participant with DESH T1-weighted MRI images on the axial (left and center) and coronal (right) planes. The CP is indicated by arrows. (A) Female (60s), MMSE score: 30/30, VS volume=0.0280, SF volume=0.0127, SHM volume=0.0360, CP volume=0.0021. (B) Male (60s), MMSE score: 25/30, VS volume=0.0406, SF volume=0.0178, SHM volume=0.0177, CP volume=0.0034. DESH-related CSF spaces and CP volumes are normalized to the total intracranial volume. *CP* choroid plexus, *DESH* disproportionately enlarged subarachnoid space hydrocephalus, *MMSE* Mini Mental State Examination, *SF* Sylvian fissure, *SHM* subarachnoid space at high convexity and midline, *VS* ventricular system.

**Figure 2.**
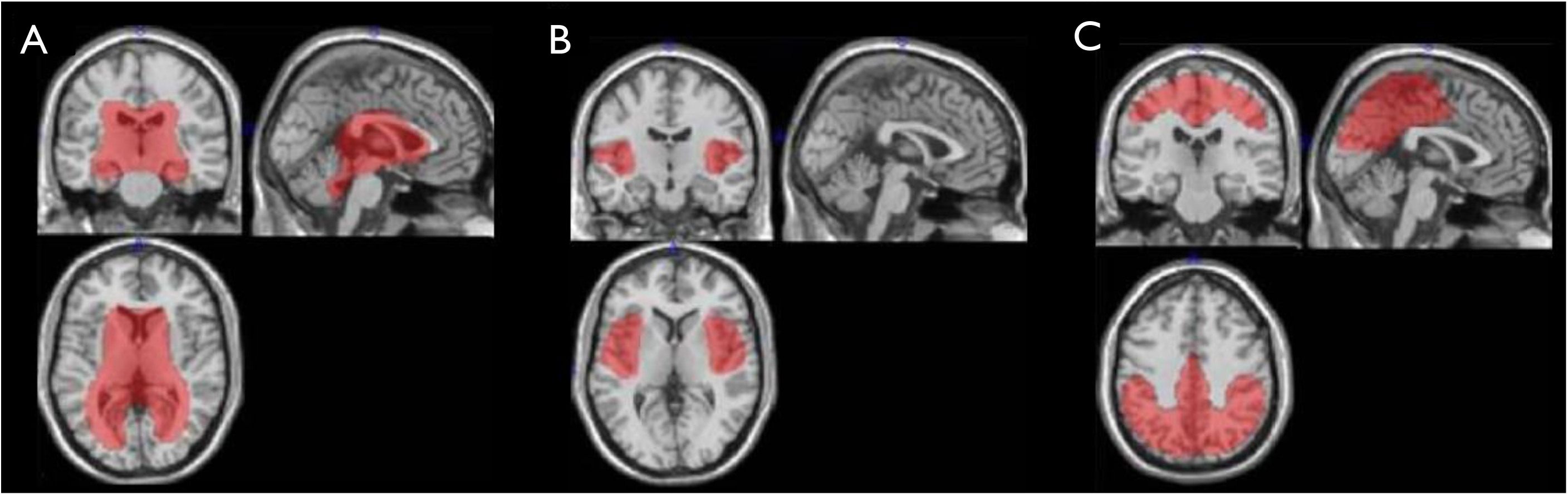
VOI template for DESH-related regions VOI templates for the CSF volumes of the VS (A), SF (B), and SHM (C). *DESH* disproportionately enlarged subarachnoid space hydrocephalus, *SF* Sylvian fissure, *SHM* subarachnoid space at high convexity and midline, *VOI* voxel of interest, *VS* ventricular system.

The choroid plexus (CP) is a richly vascularized veil-like structure located in the VS of the brain (Figure 1). [3] The CP is involved in CSF production, [4] neurogenesis, [5] removal of metabolic waste products, [6] regulation of neuroinflammation, [7] gut microbiota–immune interactions, [8] circadian rhythms [9] and CSF-mediated homeostasis of the central nervous system. [3] In recent years, evidence has been accumulating that CP is associated with the pathophysiology of Alzheimer’s disease (AD) [6,10–13] and that CP volume increases are associated with the severity of cognitive dysfunction in patients with AD. [11,12] Murine studies have suggested that CP aging is associated with cognitive decline via inflammatory cytokines. [14–16] The CP may be a potential key player in the pathophysiology of cognitive impairment in older adults; however, few studies have examined CP volume and cognitive impairment in the pre-dementia stage in community-dwelling older people. One study reported that increased CP volume is an imaging biomarker for diagnosing mild cognitive impairment (MCI) in community-dwelling older people. [17] The CP is located within the VS, and the CP volume increase may be influenced by ventricular enlargement and brain atrophy, but previous studies did not consider the volumes of both CSF and the brain parenchyma. [17] As brain atrophy and CSF volume are associated with cognitive decline in older individuals, [1] the volumes of the brain parenchyma and CSF should also be considered to investigate whether CP volume increase can be an imaging marker of cognitive dysfunction in older adults. The extent to which the CP is involved in cognitive decline in older adults compared with brain parenchymal and CSF volume remains unclear. Furthermore, the factors associated with the CP volume increase were not investigated in a large number of older people.

Therefore, this study used the data on CSF volume in DESH-related regions to investigate the following questions about the association between CP volume and cognitive decline in older adults without dementia, which have not been clarified in previous studies. Our objectives were (i) to clarify whether CP volume, independent of the volume of the brain parenchyma and CSF, can be an imaging marker of cognitive decline in older adults; (ii) to verify how strongly CP volume is associated with cognitive decline in older individuals compared with brain parenchymal and CSF volume; and (iii) to explore the factors associated with the CP volume.

## METHODS

### STUDY DESIGN AND PARTICIPANTS

This cross-sectional analysis using baseline data was part of the Japan Prospective Studies Collaboration for Aging and Dementia, which aimed to enroll approximately 10,000 community-dwelling residents aged ≥65 years from eight sites in Japan to explore genetic and environmental risk factors for dementia. [18] For this study, we enrolled 1,577 community-dwelling older residents from Arao City, Kumamoto Prefecture (southern Japan), between November 2016 and March 2017. [18] Participants were excluded if they had no or unsuitable MRI data (*n*=76 and *n*=58, respectively), missing data (*n*=23), or dementia (Online Supplemental Tables S1–3). A non-Japanese participant who had difficulty performing cognitive tests owing to language problems was excluded (*n*=1).

### PROCEDURES

Standardized approaches for questionnaires, blood tests, and dementia diagnoses were applied across all study sites, as previously described. [18] Cognitive function was assessed using the Mini-Mental State Examination (MMSE). [19] Dementia and its subtypes were diagnosed based on standard criteria, [18] and Petersen’s criteria were used to diagnose MCI. [20] Details of these diagnostic methods have been reported in a previous study. [18]

We obtained information on hypertension, diabetes mellitus (DM), dyslipidemia, atrial fibrillation, coronary artery disease, heart failure, body mass index (BMI), and history of smoking. Detailed information on the clinical data is described in a previous study. [1]

### IMAGING

A detailed description of brain MRI data acquisition and processing is provided in the Online Supplemental Material. Fluid-attenuated inversion recovery MRI was used to assess vascular diseases, (e.g., infarction and white matter hyperintensity [WMH]). Participants with large vascular lesions (e.g., cortical infarction or hemorrhage), tumors, or artifacts were excluded (Online Supplemental Table S1). Methods for the assessment of WMH, lacunar infarction, and microbleeds are described in Online Supplemental Materials.

To quantitatively assess DESH-related regions, we used an automatic volumetric segmented brain imaging system that was modified to evaluate iNPH. [21,22] We prepared voxel of interest templates for intracranial volume, VS, SF, and SHM (Figure 2), as previously described. [1,23] The details of image analysis are provided in Online Materials. Each regional volume was normalized to the total intracranial volume.

Volumetric segmentation was achieved using FreeSurfer version 5.3 (https://surfer.nmr.mgh.harvard.edu/) on CentOS6 and preprocessed according to standard procedures. [18] Using the Desikan–Killany Atlas, we measured 34 cortical regions, 7 subcortical regions, vessels (representing the perivascular space around the basal ganglia), and the CP in absolute volumes (Online Supplemental Table S4). [24] The total volumes of 34 cortical regions and 7 subcortical regions and vessel volumes were used as a covariate. Each regional volume was normalized to the total intracranial volume. All automated FreeSurfer segmentations were assessed to determine the presence of image defects.

### STATISTICAL ANALYSES

All statistical analyses were performed using SPSS version 29.0 (IBM Corp., Armonk, NY, USA). To determine whether the associations between MMSE score and the volumes of brain structures were significant when participants with MCI were excluded, we analyzed the data in two groups: all individuals (*n*=1,370) and the non-MCI group (*n*=1,144).

To investigate the association between cognitive function and CP volume, we tested the association between MMSE scores (dependent variables) and age, sex, education, hypertension, DM, dyslipidemia, atrial fibrillation, coronary artery disease, heart failure, BMI, history of smoking, MRI scanner, Fazekas score, lacunar infarction, microbleeds, perivascular space, DESH-related regions (i.e., VS, SF, and SHM), and cortical/subcortical regions (i.e., the sum of 34 cortical regions and 7 subcortical regions volumes) and CP in multiple linear regression analysis.

To verify the extent to which the volume of the intracranial structures is associated with cognitive function, we performed hierarchical multiple regression analysis using the MMSE score as the dependent variable. As the independent variables, each brain structure was separately added to the basic model, which included age, sex, education, hypertension, DM, dyslipidemia, atrial fibrillation, coronary artery disease, heart failure, BMI, history of smoking, MRI scanner, Fazekas score, lacunar infarction, microbleeds, and perivascular space. For each analysis, the standardized regression coefficient (β_STD_) and change in R^2^ (ΔR^2^) were calculated. We ranked the effects of brain structures on MMSE scores by descending Δ R^2^.

To evaluate the brain structures associated with CP volume, the hierarchical multiple regression analysis described above was performed.

To explore the CP-associated factors, we used multivariate linear regression while adjusting for the aforementioned confounders.

For all multivariate analyses, we visually inspected the normal Q-Q plots to assess the normality of the residuals. Data were examined for collinearity (variance inflation factor >10), autocorrelation (Durbin–Watson statistic values <1 or >3), and influential outliers (Cook’s distance >0.5).

For all analyses, significance was set at *P*<0.05. To correct for multiple comparisons, we used Bonferroni corrections; for example, the significance level for the comparison of 45 brain structures (41 brain regions, three DESH-related regions, and the CP) was *P*<0.0011 (0.05/45).

## RESULTS

Overall, 207 participants were excluded; thus, 1,370 participants (mean age, 73.9±6.3 years; 845 women [61.7%]) were included in the analysis. 226 participants were diagnosed with MCI. Table 1 and Online Supplemental Table S5 show the demographic and clinical characteristics of the study population.

**Table 1:** Demographic and clinical characteristics of the study participants.

|  | All participants<br>(*n*=1,370) | Non-MCI group<br>(*n*=1,144) |
| --- | --- | --- |
| Age (years) | 73.9 (6.3) | 73.1 (6.0) |
| Sex (female) | 845 (61.7%) | 727 (63.5%) |
| Education ($\leq 9$ years) | 362 (26.4%) | 266 (23.3%) |
| Hypertension | 984 (71.8%) | 813 (71.1%) |
| Diabetes mellitus | 212 (15.5%) | 174 (15.2%) |
| Dyslipidemia | 698 (50.9%) | 592 (51.7%) |
| Atrial fibrillation | 85 (6.2%) | 64 (5.6%) |
| Coronary artery disease | 72 (5.3%) | 60 (5.2%) |
| Heart failure | 19 (1.4%) | 13 (1.1%) |
| BMI | 23.5 (3.3) | 23.5 (3.2) |
| History of smoking | 355 (25.9%) | 307 (26.8%) |
| MMSE score | 27.4 (2.4) | 28.0 (2.0) |
| Imaging |  |  |
| MRI scanner (Philips) | 911 (66.5%) | 759 (66.3%) |
| Fazekas score<sup>a</sup> | 2.2 (2.1) | 2.0 (2.1) |
| Lacunar infarction | 308 (22.5%) | 236 (20.6%) |
| Microbleeds | 190 (13.9%) | 154 (13.5%) |
| Perivascular space | 0.0001 (0.0001) | 0.0001 (0.0001) |
| VS | 0.0335 (0.0089) | 0.0327 (0.0084) |
| SF | 0.0137 (0.0023) | 0.0135 (0.0021) |
| SHM | 0.0370 (0.0075) | 0.0375 (0.0071) |
| Cortical/subcortical regions<sup>b</sup> | 0.2888 (0.0229) | 0.2908 (0.0007) |
| Choroid plexus | 0.0025 (0.0004) | 0.0025 (0.0004) |

Data are presented as *n* (%) or means (standard deviations).
<sup>a</sup>The sum of periventricular and deep WMH scores, ranging from 0 to 6
<sup>b</sup>The total volumes of 34 cortical regions and 7 subcortical regions
The volumes of the perivascular space, DESH-related regions, cortical/subcortical regions, and choroid plexus were normalized to the total intracranial volume.
\*Significance level at $P < 0.05$
*BMI* body mass index, *MCI* mild cognitive impairment, *MMSE* Mini-Mental State Examination, *SF* Sylvian fissure, *SHM* subarachnoid space at the high convexity and midline, *VS* ventricular system, *WMH* white matter hyperintensity.

CP was significantly associated with MMSE scores, even after adjusting for DESH-related CSF spaces and cortical/subcortical regions (*P*=0.0065). Similar associations were observed in the non-MCI group (*P*=0.0063) (Table 2).

**Table 2:** Multivariate analysis of MMSE score.

| Variables | All participants<br>($n=1,370$) | | Participants without MCI<br>($n=1,144$) | |
| --- | --- | --- | --- | --- |
| | $\beta_{STD}$ | $P$-value | $\beta_{STD}$ | $P$-value |
| Age | -0.2160 | <0.0001* | -0.1552 | <0.0001* |
| Sex (female) | 0.1142 | 0.0008* | 0.0207 | 0.6057 |
| Education ($\leq 9$ years) | -0.1926 | <0.0001* | -0.1809 | <0.0001* |
| Hypertension | 0.0091 | 0.7319 | -0.0205 | 0.4957 |
| Diabetes mellitus | -0.0149 | 0.5593 | -0.0206 | 0.4811 |
| Dyslipidemia | 0.0133 | 0.6040 | 0.0295 | 0.3097 |
| Atrial fibrillation | -0.0263 | 0.3014 | -0.0253 | 0.3833 |
| Coronary artery disease | 0.0014 | 0.9547 | -0.0068 | 0.8165 |
| Heart failure | -0.0152 | 0.5464 | 0.0194 | 0.5011 |
| BMI | -0.0094 | 0.7162 | -0.0101 | 0.7312 |
| History of smoking | 0.0717 | 0.0263* | 0.0085 | 0.8235 |
| MRI scanner (Philips) | 0.0214 | 0.4854 | 0.0038 | 0.9127 |
| Fazekas score<sup>a</sup> | -0.0134 | 0.6760 | 0.0131 | 0.7176 |
| Lacunar infarction | -0.0090 | 0.7759 | -0.0117 | 0.7448 |
| Microbleeds | 0.0096 | 0.7171 | 0.0294 | 0.3258 |
| Perivascular space | 0.0003 | 0.9908 | 0.0011 | 0.9699 |
| VS | 0.0270 | 0.4885 | 0.0169 | 0.6994 |
| SF | -0.0046 | 0.8927 | -0.0370 | 0.3293 |
| SHM | 0.0705 | 0.0572 | 0.0080 | 0.8447 |
| Cortical/subcortical regions<sup>b</sup> | 0.0627 | 0.0787 | 0.0292 | 0.4609 |
| Choroid plexus | -0.0877 | 0.0065* | -0.1006 | 0.0063* |

\*The significance level was $P<0.05$ .
<sup>a</sup>The sum of periventricular and deep WMH scores, ranging from 0 to 6
<sup>b</sup>The total volumes of 34 cortical regions and 7 subcortical regions
BMI body mass index, MCI mild cognitive impairment, WMH white matter hyperintensity, VS ventricular system.

The results of the hierarchical multiple regression analysis of MMSE scores and brain structures are presented in Table 3. For each analysis, the top ten brain structures are shown in descending order of Δ R2. All data are shown in Online Supplemental Table S6. When analyzed in all individuals, CP and SHM were associated with MMSE scores and had βSTD coefficients of −0.1097 (*P*<0.0001) and 0.0930 (*P*=0.0004), respectively. In individuals without MCI, CP was associated with MMSE scores and had βSTD coefficients of −0.1086 (*P*=0.0006).

**Table 3:** Results of hierarchical multiple regression analysis for MMSE score with volumes of brain structures.

| All participants (*n*=1,370) |  |  |  |  |
| --- | --- | --- | --- | --- |
| Rank | Brain structure | $\beta_{STD}$ | *P*-value | $\Delta R^2$ |
| 1 | Choroid plexus | −0.1097 | <0.0001* | 0.0096 |
| 2 | SHM | 0.0930 | 0.0004* | 0.0078 |
| 3 | VS | −0.0815 | 0.0033 | 0.0054 |
| 4 | Middle temporal | 0.0832 | 0.0040 | 0.0051 |
| 5 | Supramarginal | 0.0748 | 0.0085 | 0.0043 |
| 6 | Accumbens | 0.0753 | 0.0093 | 0.0042 |
| 7 | Hippocampus | 0.0770 | 0.0110 | 0.0040 |
| 8 | Lateral occipital | 0.0717 | 0.0118 | 0.0039 |
| 9 | SF | −0.0676 | 0.0130 | 0.0038 |
| 10 | Amygdala | 0.0718 | 0.0131 | 0.0038 |

| Participants without MCI (*n*=1,144) |  |  |  |  |
| --- | --- | --- | --- | --- |
| Rank | Brain structure | $\beta_{STD}$ | *P*-value | $\Delta R^2$ |
| 1 | Choroid plexus | −0.1086 | 0.0006* | 0.0094 |
| 2 | Transverse temporal | 0.0797 | 0.0092 | 0.0054 |
| 3 | Supramarginal | 0.0741 | 0.0199 | 0.0043 |
| 4 | Superior frontal | 0.0738 | 0.0290 | 0.0038 |
| 5 | SF | −0.0650 | 0.0346 | 0.0036 |
| 6 | Accumbens | 0.0648 | 0.0473 | 0.0031 |
| 7 | VS | −0.0615 | 0.0481 | 0.0031 |
| 8 | SHM | 0.0529 | 0.0746 | 0.0025 |
| 9 | Banks sts | 0.0569 | 0.0826 | 0.0024 |
| 10 | Precentral | 0.0654 | 0.0834 | 0.0024 |

As the independent variables, each brain structure was separately added to the basic model, which included age, sex, education, hypertension, DM, dyslipidemia, atrial fibrillation, coronary artery disease, heart failure, BMI, history of smoking, MRI scanner, Fazekas score, lacunar infarction, microbleeds, and perivascular space. For each analysis, the standardized regression coefficient ( $\beta_{STD}$ ) and change in $R^2$ ( $\Delta R^2$ ) were calculated. The top 10 brain structures are shown in descending order of $\Delta R^2$ .
\*The significance level was $P < 0.0011$ to correct for 45 modeling analyses.
*BMI* body mass index, *MCI* mild cognitive impairment, *MMSE* Mini-Mental State Examination, *SF* Sylvian fissure, *SHM* subarachnoid space at high convexity and midline, *sts* superior temporal sulcus, *VS* ventricular system

The hierarchical multiple regression analysis results of CP volume with volumes of other brain structures are presented in Online Supplemental Table S7. Forty-four brain structures were shown in descending order of Δ R2. CP volumes were associated with 32 and 24 regions in the entire study population and the non-MCI group, respectively.

In the multivariate analysis of CP volume, VS, a history of smoking, enlarged perivascular space, a higher Fazekas score, age, BMI, DM, absence of dyslipidemia, and absence of atrial fibrillation were associated with larger CP volumes (Table 4). Similar associations were observed in the non-MCI group.

**Table 4:** Multivariate analysis of choroid plexus volume.

|  | All participants<br>(*n*=1,370) |  | Participants without MCI<br>(*n*=1,144) |  |
| --- | --- | --- | --- | --- |
| Variables | $\beta_{STD}$ | *P*-value | $\beta_{STD}$ | *P*-value |
| Age | 0.0710 | 0.0084* | 0.0764 | 0.0071* |
| Female sex | 0.0108 | 0.7084 | 0.0217 | 0.5042 |
| Education ($\leq 9$ years) | -0.0278 | 0.2096 | -0.0020 | 0.9351 |
| Hypertension | 0.0372 | 0.0973 | 0.0384 | 0.1162 |
| Diabetes mellitus | 0.0508 | 0.0188* | 0.0569 | 0.0162* |
| Dyslipidemia | -0.0463 | 0.0325* | -0.0411 | 0.0807 |
| Atrial fibrillation | -0.0618 | 0.0041* | -0.0537 | 0.0224* |
| Coronary artery disease | -0.0212 | 0.3263 | -0.0241 | 0.3089 |
| Heart failure | 0.0181 | 0.3954 | 0.0074 | 0.7511 |
| BMI | 0.0626 | 0.0041* | 0.0688 | 0.0039* |
| History of smoking | 0.1413 | <0.0001* | 0.1431 | <0.0001* |
| MRI scanner (Philips) | -0.1002 | 0.0001* | -0.0734 | 0.0090* |
| Fazekas score<sup>a</sup> | 0.1098 | 0.0001* | 0.1251 | <0.0001* |
| Lacunar infarction | -0.0410 | 0.1267 | -0.0610 | 0.0371* |
| Microbleeds | 0.0391 | 0.0802 | 0.0303 | 0.2121 |
| Perivascular space | 0.0887 | <0.0001* | 0.0795 | 0.0008* |
| VS | 0.4685 | <0.0001* | 0.4790 | <0.0001* |
| SF | 0.0313 | 0.2762 | 0.0693 | 0.0243* |
| SHM | -0.0248 | 0.4292 | -0.0079 | 0.8131 |
| Cortical/subcortical regions<sup>b</sup> | -0.0130 | 0.6661 | 0.0242 | 0.4511 |

\*The significance level was set at $P < 0.05$ .
<sup>a</sup>The sum of periventricular and deep WMH scores, ranging from 0 to 6
<sup>b</sup>The total volumes of 34 cortical regions and 7 subcortical regions
*BMI* body mass index, *MCI* mild cognitive impairment, *WMH* white matter hyperintensity

We did not find any violation of linear regression assumptions, and none of the analyses were influenced by an influential outlier.

## DISCUSSION

This study examined the association between CP volume and cognitive impairment in community-dwelling older adults without dementia. Increased CP volumes were significantly associated with lower MMSE scores, even after adjusting for DESH-related CSF spaces and cortical or subcortical region volumes. CP volumes were more strongly associated with MMSE scores than the volumes of DESH-related CSF spaces and brain parenchyma. Furthermore, these results were similar in the non-MCI group. Previously, we reported that CSF volumes in DESH-related regions, particularly in SHM, were associated with cognitive decline in older individuals. [1] However, the present results suggest that CP volume can be a more sensitive imaging marker of cognitive decline in older individuals than CSF volumes in DESH-related regions and the brain parenchyma. To the best of our knowledge, this is the first study to examine the association between CP volume increase and cognitive decline in older individuals using volume data from both the CSF and brain parenchyma in a large number of community-dwelling older individuals.

Why can CP volume be a sensitive imaging marker of cognitive impairment in older adults? The brain structures most strongly associated with CP volume were CSF spaces related to DESH (i.e., VS, SHM, and SF) and AD-related regions (accumbens, hippocampus, amygdala, inferior temporal, posterior cingulate, supramarginal, lateral orbitofrontal). [25,26] Larger CP volumes indicate larger VS and SF volumes and smaller SHM volumes, suggesting that the brain morphology of older individuals is more “DESH-like.” Moreover, the larger the CP volume, the smaller the volume of AD-related regions, indicating that the brain morphology of older adults is more “AD-like.” These results suggest that the CP may be involved in the pathophysiology of both iNPH (i.e., “CSF dynamics disorders” [27]) and AD. Similar results were obtained when the MCI group was excluded, suggesting that the CP is continuously involved in the pathophysiology of CSF dynamic disorders and AD from the pre-MCI to the MCI stage. To the best of our knowledge, no prior studies have investigated CP volumes in patients with CSF dynamic disorders, but many studies have investigated the possibility that CP is closely related to AD development. [6,10–13] As CSF dynamics disorders and AD cause cognitive decline in older individuals, these conditions may partly explain the association between CP volume increase and cognitive decline in older adults. However, in this study, the increased CP volumes were significantly associated with lower MMSE scores, even after adjusting for DESH-related CSF spaces and cortical or subcortical region volumes. These results suggest that increases in CP are associated with cognitive decline owing to factors other than CSF dynamics dysfunction or neurodegeneration. What other factors influence the association between increases in CP and cognitive decline in older adults? At present, we can only speculate based on the results of previous studies. In recent years, evidence has been accumulating that increased CP volume is associated with the pathophysiology of neurological and psychiatric disorders via neuroinflammation. [10,28,29] A positive correlation between CP volume and plasma glial fibrillary acidic protein levels, a marker of neuroinflammation, has been reported in patients with AD. [10] In multiple sclerosis, increased neuroinflammation leads to increased CP volume, which can be an imaging marker of dysfunction in both humans and mice. [28] Among patients with schizophrenia, plasma interleukin (IL)-6 levels are higher in patients with larger CP volume. [29] Neuroinflammation may be involved in the mechanism of CP volume increase; however, to the best of our knowledge, no prior studies reported an association between CP aging, an exacerbation of the age-related neuroinflammatory response, and cognitive dysfunction in humans. Murine studies have suggested that CP aging is associated with cognitive decline via inflammatory cytokines. [14–16] Type I interferon signaling in the CP is associated with cognitive decline and reduced hippocampal neurogenesis in older mice, [14] and interferon-β overexpression in the CP of younger mice induces an aging-like microglial phenotype and cognitive dysfunction. [15] In the CP of older mice, T-helper type 2-derived cytokine IL-4 levels are elevated and interferon-γ levels are decreased, and this local cytokine shift triggers the production of the chemokine CCL11, which is associated with cognitive dysfunction. [16] The blood–CSF barrier of the CP plays an important role in the transmission of inflammatory signals from the periphery to the central nervous system. [3,7] The age-related disruption of the blood–CSF barrier may cause an inflammatory response in the brain, [30] which in turn may lead to cognitive decline in older individuals. In the present study, these associations could not be verified because inflammatory cytokine levels were not measured. Further studies are required to determine the association between CP-dependent inflammatory responses in the brain and cognitive decline in older adults.

Factors associated with increased CP volume include age, WMH, enlarged perivascular spaces, BMI, history of smoking, and DM. CP volume increased with age, independent of the volume of the CSF and brain parenchyma. This suggests that the age-related increase in the volume of the CP is not secondary to brain atrophy or ventricular enlargement. Pathological age-related changes in the CP have been reported, including a thickening of epithelial basement membranes, irregular fibrosis of the stroma, and accumulation of calcification. [31,32] Chronic inflammation appears to be closely linked to aging. [33] Neuroinflammation may be associated with CP volume increases in neurological and psychiatric disorders. [10,28,29] However, the mechanism of age-related chronic inflammation and CP volume increase remains unclear. Further studies on aging processes in CP are required. Associations with increased CP volume, WMH enlargement, and impaired glymphatic function have been reported. [34] The volume of the perivascular space, a structure of the glymphatic system involved in excreting metabolites, [35] is associated with the CP volume. Inflammation may reduce glymphatic function, [36] suggesting that increased CP volume and glymphatic dysfunction may be associated via an inflammatory response. A previous study reported that BMI was associated with increased CP volume, suggesting that obesity-induced inflammation leads to increased CP volume. [37] To the best of our knowledge, a history of smoking or DM has not been previously associated with increased CP volume, but inflammation caused by smoking [38] or DM [39] may be associated with increased CP volume. Smoking cessation and DM prevention or treatment may reduce CP volume increases; however, this needs to be validated in future studies. Previous studies and our present results suggest that inflammation may play a role in increases in CP volume; however, the mechanisms are not fully understood; thus, further studies are required. Elucidating the mechanisms of CP volume increase can lead to early intervention for cognitive decline in older adults. Our results emphasize that more attention should be paid to CP volume increase to maintain cognitive function in older individuals.

The strength of this study is that it analyzed the association between CP volume and cognitive function in over 1,300 community-dwelling older adults using CSF and brain parenchymal volume data. To the best of our knowledge, no previous study has directly measured CSF volumes in the three DESH-related regions in a large number of older individuals, making the present results novel and unique.

This study had some limitations. First, the CP is located in the CSF within the VS, raising the question of whether its volume can be accurately measured. Manual tracing is the most accurate segmentation method but impractical for analyses of large numbers of images. Several studies have used the FreeSurfer software to calculate CP volume by automated CP segmentation using three-dimensional T1-weighted images. [10–12,17,29,40] CP volumes measured using FreeSurfer version 5.3 were larger in patients with AD than in healthy individuals, and this result did not change in analyses using manually measured volumes. [12] Thus, the CP volumes measured in this study might be sufficiently accurate to analyze a large number of cases. Second, this cross-sectional analysis was unable to identify causal associations; thus, future longitudinal studies are required. In particular, the association between increased CP volumes and cognitive impairment in older individuals requires further longitudinal verification. Finally, an MRI was performed using two different devices. To eliminate bias, the scanner type was included in multivariate regression analysis as a covariate.

## CONCLUSIONS

In community-dwelling older individuals without dementia, CP volume is associated with cognitive decline independent of the volumes of brain parenchyma and CSF and may be one of the most sensitive imaging markers of cognitive decline. Our findings emphasize the importance of investigating the CP volume increase to maintain cognitive function in older individuals. Therefore, further longitudinal studies are required.

## Data Availability

The raw data are not openly available to protect the confidentiality of the participants and to comply with the terms of participant consent. Requests related to the raw data should be addressed to the corresponding author.

## ACKNOWLEDGMENTS

The authors are deeply grateful to all the participants and staff involved in this study. The authors thank the staff of Arao City, Arao Municipal Hospital, and Social Insurance Omuta Tenryo Hospital for their support. The authors also thank the members of the Department of Neuropsychiatry at Kumamoto University and Editage (www.editage.com) for the English language editing.

## ETHICS APPROVAL AND CONSENT TO PARTICIPATE

The Research Ethics Committee of Kumamoto University (Kumamoto, Japan [approval number: GENOME-333]) approved this study. All participants provided written informed consent prior to data collection in accordance with the Declaration of Helsinki.

## AUTHORS’ CONTRIBUTIONS

YH contributed to the study design, data collection, data analysis, and manuscript writing and revision. MH, RF, TI, NT, AK, KH, YM, KY, and SY contributed to the study design, data collection, data analysis, and manuscript revision. TS and KI contributed to the study design, data analysis, and manuscript revision. MI contributed to the study design, data collection, and manuscript revision. NK, SB, and MT contributed to the study design and manuscript revision. All authors had the final responsibility for the decision to submit the manuscript for publication.

## REFERENCES

1 Hidaka Y, Hashimoto M, Suehiro T, et al. Impact of age on the cerebrospinal fluid spaces: high-convexity and medial subarachnoid spaces decrease with age. Fluids Barriers CNS Oct 28 2022;19:82. doi:10.1186/s12987-022-00381-5.

2 Hashimoto M, Ishikawa M, Mori E et al. Diagnosis of idiopathic normal pressure hydrocephalus is supported by MRI-based scheme: a prospective cohort study. Cerebrospinal Fluid Res Oct 31 2010;7:18. doi:10.1186/1743-8454-7-18.

3 Ghersi-Egea JF, Strazielle N, Catala M et al. Molecular anatomy and functions of the choroidal blood-cerebrospinal fluid barrier in health and disease. Acta Neuropathol Mar 2018;135:337–61. doi:10.1007/s00401-018-1807-1.

4 Lun MP, Monuki ES, Lehtinen MK. Development and functions of the choroid plexus-cerebrospinal fluid system. Nat Rev Neurosci Aug 2015;16:445–57. doi:10.1038/nrn3921.

5 Silva-Vargas V, Maldonado-Soto AR, Mizrak D et al. Age-dependent niche signals from the choroid plexus regulate adult neural stem cells. Cell Stem Cell Nov 3 2016;19:643–52. doi:10.1016/j.stem.2016.06.013.

6 Duarte AC, Furtado A, Hrynchak MV, et al. Age, sex hormones, and circadian rhythm regulate the expression of amyloid-beta scavengers at the choroid plexus. Int J Mol Sci Sep 17 2020;21.

7 Solár P, Zamani A, Kubíčková L et al. Choroid plexus and the blood-cerebrospinal fluid barrier in disease. Fluids Barriers CNS May 6 2020;17:35. doi:10.1186/s12987-020-00196-2.

8 Xie J, Bruggeman A, De Nolf C, et al. Gut microbiota regulates blood-cerebrospinal fluid barrier function and Aβ pathology. EMBO J Sep 4 2023;42:e111515. doi:10.15252/embj.2022111515.

9 Myung J, Schmal C, Hong S, et al. The choroid plexus is an important circadian clock component. Nat Commun Mar 14 2018;9:1062. doi:10.1038/s41467-018-03507-2.

10 Bouhrara M, Walker KA, R Alisch JS, et al. Association of plasma Markers of Alzheimer’s disease, neurodegeneration, and neuroinflammation with the choroid plexus integrity in aging. Aging Dis Jan 8 2024. doi:10.14336/AD.2023.1226.

11 Choi JD, Moon Y, Kim HJ et al. Choroid plexus volume and permeability at brain MRI within the Alzheimer disease clinical spectrum. Radiology Sep 2022;304:635–45. doi:10.1148/radiol.212400.

12 Čarna M, Onyango IG, Katina S, et al. Pathogenesis of Alzheimer’s disease: involvement of the choroid plexus. Alzheimers Dement Aug 2023;19:3537–54. doi:10.1002/alz.12970.

13 Jeong SH, Park CJ, Cha J, et al. Choroid plexus volume, amyloid burden, and cognition in the Alzheimer’s disease continuum. Aging Dis Jan 23 2024. doi:10.14336/AD.2024.0118.

14 Baruch K, Deczkowska A, David E, et al. Aging. Aging-induced type I interferon response at the choroid plexus negatively affects brain function. Science Oct 3 2014;346:89–93. doi:10.1126/science.1252945.

15 Deczkowska A, Matcovitch-Natan O, Tsitsou-Kampeli A, et al. Mef2C restrains microglial inflammatory response and is lost in brain ageing in an IFN-I-dependent manner. Nat Commun Sep 28 2017;8:717. doi:10.1038/s41467-017-00769-0.

16 Baruch K, Ron-Harel N, Gal H, et al. CNS-specific immunity at the choroid plexus shifts toward destructive Th2 inflammation in brain aging. Proc Natl Acad Sci U S A Feb 5 2013;110:2264–9. doi:10.1073/pnas.1211270110.

17 Umemura Y, Watanabe K, Kasai S, et al. Choroid plexus enlargement in mild cognitive impairment on MRI: a large cohort study. Eur Radiol Jan 15 2024;34:5297–304. doi:10.1007/s00330-023-10572-9.

18 Ninomiya T, Nakaji S, Maeda T, et al. Study design and baseline characteristics of a population-based prospective cohort study of dementia in Japan: the Japan Prospective Studies Collaboration for Aging and Dementia (JPSC-AD). Environ Health Prev Med Oct 31 2020;25:64. doi:10.1186/s12199-020-00903-3.

19 Folstein MF, Folstein SE, McHugh PR. ‘Mini-mental state’. A practical method for grading the cognitive state of patients for the clinician. J Psychiatr Res Nov 1975;12:189–98. doi:10.1016/0022-3956(75)90026-6.

20 Petersen RC, Stevens JC, Ganguli M et al. Practice parameter: early detection of dementia: mild cognitive impairment (an evidence-based review). Report of the Quality Standards Subcommittee of the American Academy of Neurology. Neurology May 8 2001;56:1133–42. doi:10.1212/wnl.56.9.1133.

21 Ishii K, Soma T, Kono AK, et al. Automatic volumetric measurement of segmented brain structures on magnetic resonance imaging. Radiat Med Jul 2006;24:422–30. doi:10.1007/s11604-006-0048-8.

22 Ishii K, Soma T, Shimada K et al. Automatic volumetry of the cerebrospinal fluid space in idiopathic normal pressure hydrocephalus. Dement Geriatr Cogn Dis Extra Jan 2013;3:489–96. doi:10.1159/000357329.

23 Suehiro T, Kazui H, Kanemoto H, et al. Changes in brain morphology in patients in the preclinical stage of idiopathic normal pressure hydrocephalus. Psychogeriatrics Nov 2019;19:557–65. doi:10.1111/psyg.12445.

24 Desikan RS, Ségonne F, Fischl B, et al. An automated labeling system for subdividing the human cerebral cortex on MRI scans into gyral based regions of interest. Neuroimage Jul 1 2006;31:968–80. doi:10.1016/j.neuroimage.2006.01.021.

25 Tang X, Guo Z, Chen G, et al. A multimodal meta-analytical evidence of functional and structural brain abnormalities across Alzheimer’s disease spectrum. Ageing Res Rev Mar 2024;95:102240. doi:10.1016/j.arr.2024.102240.

26 Shulman D, Dubnov S, Zorbaz T, et al. Sex-specific declines in cholinergic-targeting tRNA fragments in the nucleus accumbens in Alzheimer’s disease. Alzheimers Dement Nov 2023;19:5159–72. doi:10.1002/alz.13095.

27 Graff-Radford J, Gunter JL, Jones DT, et al. Cerebrospinal fluid dynamics disorders: relationship to Alzheimer biomarkers and cognition. Neurology Dec 10 2019;93:e2237–46-e2246. doi:10.1212/WNL.0000000000008616.

28 Fleischer V, Gonzalez-Escamilla G, Ciolac D, et al. Translational value of choroid plexus imaging for tracking neuroinflammation in mice and humans. Proc Natl Acad Sci U S A Sep 7 2021;118.

29 Lizano P, Lutz O, Ling G, et al. Association of choroid plexus enlargement with cognitive, inflammatory, and structural phenotypes across the psychosis spectrum. Am J Psychiatry Jul 1 2019;176:564–72. doi:10.1176/appi.ajp.2019.18070825.

30 Erickson MA, Banks WA. Age-Associated Changes in the Immune System and BloodLBrain Barrier Functions. Int J Mol Sci. Age: Associated Apr 2 2019;20.

31 Serot JM, Foliguet B, Béné MC et al. Choroid plexus and ageing in rats: a morphometric and ultrastructural study. Eur J Neurosci Sep 2001;14:794–8. doi:10.1046/j.0953-816x.2001.01693.x.

32 Yalcin A, Ceylan M, Bayraktutan OF et al. Age and gender related prevalence of intracranial calcifications in CT imaging; data from 12,000 healthy subjects. J Chem Neuroanat Dec 2016;78:20–4. doi:10.1016/j.jchemneu.2016.07.008.

33 Li X, Li C, Zhang W et al. Inflammation and aging: signaling pathways and intervention therapies. Signal Transduct Target Ther Jun 8 2023;8:239. doi:10.1038/s41392-023-01502-8.

34 Li Y, Zhou Y, Zhong W, et al. Choroid plexus enlargement exacerbates white matter hyperintensity growth through glymphatic impairment. Ann Neurol Jul 2023;94:182–95. doi:10.1002/ana.26648.

35 Iliff JJ, Wang M, Liao Y, et al. A paravascular pathway facilitates CSF flow through the brain parenchyma and the clearance of interstitial solutes, including amyloid β. Sci Transl Med Aug 15 2012;4:147ra111. doi:10.1126/scitranslmed.3003748.

36 Cai Y, Zhang Y, Leng S, et al. The relationship between inflammation, impaired glymphatic system, and neurodegenerative disorders: A vicious cycle. Neurobiol Dis Mar 2024;192:106426. doi:10.1016/j.nbd.2024.106426.

37 Alisch JSR, Egan JM, Bouhrara M. Differences in the choroid plexus volume and microstructure are associated with body adiposity. Front Endocrinol (Lausanne) 2022;13:984929. doi:10.3389/fendo.2022.984929.

38 Liu Y, Li H, Wang J, et al. Association of cigarette smoking with cerebrospinal fluid biomarkers of neurodegeneration, neuroinflammation, and oxidation. JAMA Netw Open Oct 1 2020;3:e2018777. doi:10.1001/jamanetworkopen.2020.18777.

39 Donath MY, Shoelson SE. Type 2 diabetes as an inflammatory disease. Nat Rev Immunol Feb 2011;11:98–107. doi:10.1038/nri2925.

40 Assogna M, Premi E, Gazzina S, et al. Association of choroid plexus volume with serum biomarkers, clinical features, and disease severity in patients with frontotemporal lobar degeneration spectrum. Neurology Sep 19 2023;101:e1218–30-e1230. doi:10.1212/WNL.0000000000207600.

