## Supplementary material for "Association between the choroid plexus volume and cognitive function in community-dwelling older adults without dementia: A population-based cross-sectional analysis"

**Supplemental Methods**

**Imaging**

Brain magnetic resonance imaging (MRI) was conducted at the Arao Municipal Hospital (Kumamoto, Japan) and Omuta Tenryo Hospital (Fukuoka, Japan) using the 1.5-Tesla Ingenia CX dual scanner (Philips Healthcare, Best, Netherlands) or the 1.5-Tesla Signa HDxt Ver.23 scanner (GE Healthcare, Milwaukee, WI, USA). The Philips MRI scanning protocol consisted of a three-dimensional (3D) T1-weighted sequence (repetition time=8.6 ms, echo time=4.0 ms, flip angle=9°, matrix=192×192, slice thickness=1.2 mm), a 3D T2-weighted sequence (repetition time=5082.3 ms, echo time=100.0 ms, flip angle=90°, matrix=356×248, slice thickness=5.0 mm), a 3D fluid-attenuated inversion recovery (FLAIR) sequence (repetition time=11000.0 ms, echo time=120.0 ms, flip angle=90°, matrix=288×203, slice thickness=5.0 mm), and a susceptibility-weighted imaging (SWI) sequence (repetition time=78.4 ms, echo time=41.4 ms, flip angle=20°, matrix=88×272, slice thickness=2.4 mm). The GE Signa MRI scanning protocol consisted of a 3D T1-weighted sequence (repetition time=8.3 ms, echo time=3.4 ms, flip angle=8°, matrix=192 × 192, slice thickness=1.2 mm), a 3D T2-weighted sequence (repetition time=4517.0 ms, echo time=92.6 ms, flip angle=90°, matrix=352 × 224, slice thickness=5.0 mm), a 3D FLAIR sequence (repetition time=10000.0 ms, echo time=149.2 ms, flip angle=90°, matrix=288 × 193, slice thickness=5.0 mm), and a 3D T2-Star weighted angiography sequence (repetition time=75.2 ms, echo time=57.9 ms, flip angle=20°, matrix=320×200, slice thickness=3.0 mm).

Three-dimensional T1-weighted images were acquired according to the Alzheimer’s Disease Neuroimaging Initiative protocol. ^1^ FLAIR MRI was used to assess vascular diseases, e.g., infarction and white matter hyperintensity (WMH). Participants with large vascular lesions (e.g., cortical infarction or hemorrhage), tumors, or artifacts were excluded. The degree of white matter hyperintensity (WMH) load was visually rated on axial FLAIR images using the Fazekas scale (grade 1: punctate, grade 2: early confluent, or grade 3: confluent) in periventricular and deep white matter (WM) regions. ^2^ The sum of periventricular and deep WMH scores (range 0–6) was used for analysis. Lacunar infarction was defined as at least one cerebrospinal fluid (CSF)-like hypointensity with a diameter >2 mm surrounded by a hyperintense rim on T2 FLAIR. Likewise, microbleeds were assessed using SWI or T2-Star software. The Fazekas scale score, lacunar infarction, and microbleeds were considered covariates. All brain images were assessed by one neuroradiologist (N.T.) and two neuropsychiatrists (Y.H. and M.H.) blinded to the clinical data. Each evaluator assessed all brain images separately. In discordant cases, ratings were consensually determined.

To quantitatively assess disproportionately enlarged subarachnoid space hydrocephalus (DESH)-related regions, we utilized an automatic volumetric segmented brain imaging system that was modified to evaluate idiopathic normal-pressure hydrocephalus. ^3,4^ We prepared voxel of interest (VOI) templates for intracranial volume, ventricular system (VS), Sylvian fissure (SF), and subarachnoid space at the high convexity and midline (SHM) (figure 2), as previously described. ^5^ Each regional VOI template was generated using a digital phantom of the Simulated Brain Database (https://www.bic.mni.mcgill.ca/brainweb/) according to the standard Montreal Neurological Institute space with the contours of each structure manually delineated. The SHM VOI template was manually produced based on the results of a previous study on voxel-based morphometry in patients and normal controls. ^6^ For this process, the MRI of each participant was segmented into the gray matter (GM), WM, and CSF using the SPM8 segmentation program (https://www.fil.ion.ucl.ac.uk/spm/software/spm8/). The GM template derived from the Simulated Brain Database was spatially transformed into a GM image for each participant, and a normalization parameter was determined using SPM8 and the Diffeomorphic Anatomical Registration Through Exponentiated Lie Algebra technique. Using this normalization parameter, which functions like the reverse parameter generated during anatomical normalization to a standard brain, the intracranial volume and VS, SF, and SHM VOI templates were transformed into each participant’s space. The intracranial volume was adjusted using images derived from segmented GM, WM, and CSF images. Segmented GM (WM) images were derived by calculating GM (WM) areas with voxels from the intracranial volume VOI template. The CSF volumes of the VS, SF, and SHM were calculated using individual transformed VS, SF, and SHM subarachnoid space VOI templates. Each regional volume was normalized to the total intracranial volume.

1.	Jack CR, Jr., Bernstein MA, Fox NC, et al. The Alzheimer's Disease Neuroimaging Initiative (ADNI): MRI methods. *J Magn Reson Imaging*. Apr 2008;27(4):685-91. doi:10.1002/jmri.21049

2.	Fazekas F, Chawluk JB, Alavi A, Hurtig HI, Zimmerman RA. MR signal abnormalities at 1.5 T in Alzheimer's dementia and normal aging. *AJR Am J Roentgenol*. Aug 1987;149(2):351-6. doi:10.2214/ajr.149.2.351

3.	Ishii K, Soma T, Kono AK, et al. Automatic volumetric measurement of segmented brain structures on magnetic resonance imaging. *Radiat Med*. Jul 2006;24(6):422-30. doi:10.1007/s11604-006-0048-8

4.	Ishii K, Soma T, Shimada K, Oda H, Terashima A, Kawasaki R. Automatic volumetry of the cerebrospinal fluid space in idiopathic normal pressure hydrocephalus. *Dement Geriatr Cogn Dis Extra*. Jan 2013;3(1):489-96. doi:10.1159/000357329

5.	Suehiro T, Kazui H, Kanemoto H, et al. Changes in brain morphology in patients in the preclinical stage of idiopathic normal pressure hydrocephalus. *Psychogeriatrics*. Nov 2019;19(6):557-565. doi:10.1111/psyg.12445

6.	Ishii K, Kawaguchi T, Shimada K, et al. Voxel-based analysis of gray matter and CSF space in idiopathic normal pressure hydrocephalus. *Dement Geriatr Cogn Disord*. 2008;25(4):329-35. doi:10.1159/000119521

| **Table S1: The list of participants with unsuitable MRI for quantitative analysis** | | |
| --- | --- | --- |
| **age** | **sex** | **Reason for omission** |
| 60s | Male | Right and left temporal lobe contusion |
| 60s | Male | Right putamen and corona radiata infarction |
| 60s | Male | Left putaminal hemorrhage |
| 60s | Female | Frontal lobe infarction |
| 60s | Female | Right putaminal infarction |
| 60s | Male | Left putaminal infarction |
| 60s | Female | Left frontal lobe hemorrhage |
| 60s | Female | Metal artifact, post clipping for aneurysm |
| 60s | Male | Right temporo-occipital infarction |
| 60s | Male | Left frontal lobe infarction |
| 60s | Male | Metal artifact (right middle cerebral artery) |
| 60s | Male | Right occipital lobe, right putamen and left frontal lobe infarction |
| 70s | Male | Right and left temporal lobe infarction |
| 70s | Female | Right corona radiata infarction |
| 70s | Female | Skull base tumor |
| 70s | Female | Right frontal lobe infarction |
| 70s | Female | Left cerebellum infarction |
| 70s | Male | Left frontal lobe and right occipital lobe infarction |
| 70s | Female | Right putamen and corona radiata infarction |
| 70s | Male | Meningioma |
| 70s | Female | Left frontal lobe contusion |
| 70s | Male | Right putaminal hemorrhage |
| 70s | Female | Right temporal lobe infarction |
| 70s | Female | Metal artifact (post clipping for aneurysm), cerebellum infarction |
| 70s | Male | Right parietal lobe infarction |
| 70s | Male | Right frontal lobe contusion |
| 70s | Female | Metal artifact (right middle cerebral artery) |
| 70s | Male | Left frontal lobe infarction |
| 70s | Male | Right occipital lobe infarction, right putaminal hemorrhage |
| 70s | Female | Left frontal and occipital lobe infarction |
| 70s | Female | Left parietal lobe infarction |
| 70s | Female | Right putaminal hemorrhage |
| 70s | Male | Right putaminal infarction |
| 70s | Male | Left putaminal infarction |
| 70s | Female | Right frontal and temporal lobe contusion |
| 70s | Male | Left occipital lobe infarction |
| 70s | Male | Right temporal and parietal lobe infarction |
| 70s | Male | Left temporo-occipital infarction |
| 80s | Female | Metal artifact |
| 80s | Male | Left frontal lobe infarction |
| 80s | Male | Right occipital lobe and left frontal lobe infarction |
| 80s | Male | Left putaminal infarction |
| 80s | Female | Left putaminal hemorrhage |
| 80s | Female | Left middle cerebral artery territory infarction |
| 80s | Male | Left occipital lobe infarction |
| 80s | Female | Right putaminal infarction, Right temporo-occipital infarction |
| 80s | Male | Left temporo-occipital infarction |
| 80s | Female | Right frontal lobe contusion |
| 80s | Female | Right frontal lobe contusion |
| 80s | Female | Left subdural hygroma |
| 80s | Female | Left frontal lobe contusion and infarction |
| 80s | Male | Motion artifact |
| 80s | Female | Metal artifact, post clipping for aneurysm |
| 80s | Male | Right and left thalamic hemorrhage |
| 90s | Male | Left frontal lobe contusion |
| 90s | Male | Right and left subdural hygroma |
| 90s | Male | Right pons infarction |
| 90s | Female | Artifact (shunt tube) |

| **Table S2: Proportion of missing data for variables recorded in this study** | |
| --- | --- |
| **Variables** | **Number of missing data** |
| Hypertension | 4 (0.26%) |
| Diabetes Mellitus | 4 (0.26%) |
| Dyslipidemia | 9 (0.60%) |
| Atrial fibrillation | 2 (0.13%) |
| Heart failure | 2 (0.13%) |
| History of smoking | 6 (0.40%) |
| BMI | 2 (0.13%) |
| MMSE | 3 (0.20%) |

| **Table S3: Details of dementia subtype** | | |
| --- | --- | --- |
| **Dementia subtype** | **Number of participants** | **Frequency (%)** |
| AD (isolated) | 47 | 81.0 |
| VaD (isolated) | 5 | 8.6 |
| DLB (isolated) | 3 | 5.2 |
| iNPH (isolated) | 1 | 1.7 |
| Mixed type  AD+iNPH  VaD+CO toxication | 2 | 3.4 |
|  | 1 | - |
|  | 1 | - |
| AD=Alzheimer's disease; VaD=Vascular dementia; DLB=Dementia with Lewy bodies; iNPH=Idiopathic normal pressure hydrocephalus; CO=Carbon monoxide | | |

**Table S4:** **The list of brain structures whose volumes were calculated using FreeSurfer**

Frontal lobe

Caudal anterior cingulate

Caudal middle frontal

Lateral orbitofrontal

Medial orbitofrontal

Paracentral

Pars opercularis

Pars orbitalis

Pars triangularis

Precentral

Rostral anterior cingulate

Rostral middle frontal

Superior frontal

Frontal pole

Temporal lobe

Banks sts

Entorhinal

Fusiform

Inferior temporal

Middle temporal

Parahippocampal

Superior temporal

Temporal pole

Transverse temporal

Parietal lobe

Inferior parietal

Isthmus cingulate

Postcentral

Posterior cingulate

Precuneus

Superior parietal

Supramarginal

Occipital lobe

Cuneus

Lateral occipital

Lingual

Pericalcarine

Insula

Subcortical

Accumbens

Amygdala

Caudate

Hippocampus

Putamen

Pallidum

Thalamus

Choroid plexus

Vessels

**Table S5: Demographic and clinical characteristics of the MCI group**

|  | **MCI group (*n*=226)** |
| --- | --- |
| Age, years | 77.9 (6.4) |
| Female sex | 118 (52.2%) |
| Education (≤9 years) | 96 (42.5%) |
| Hypertension | 171 (75.7%) |
| Diabetes mellitus | 38 (16.8%) |
| Dyslipidemia | 106 (46.9%) |
| Atrial fibrillation | 21 (9.3%) |
| Coronary artery disease | 12 (5.3%) |
| Heart failure | 6 (2.7%) |
| BMI | 23.5 (3.5) |
| History of smoking | 48 (21.2%) |
| MMSE score | 24.5 (2.5) |
| Imaging |  |
| MRI scanner (Philips) | 152 (67.3%) |
| Fazekas score^a^ | 2.9 (2.3) |
| Lacunar infarction | 72 (31.9%) |
| Microbleeds | 36 (15.9%) |
| Perivascular space | 0.0001 (0.0001) |
| VS | 0.0378 (0.0101) |
| SF | 0.0145 (0.0027) |
| SHM | 0.0345 (0.0085) |
| Cortical/subcortical regions^b^ | 0.2788 (0.0016) |
| Choroid plexus | 0.0027 (0.0004) |

Data are presented as *n* (%) or means (standard deviations).

^a^The sum of periventricular and deep WMH scores, ranging from 0 to 6

^b^The total volumes of 34 cortical regions and 7 subcortical regions

The volumes of the perivascular space, DESH-related regions, cortical/subcortical regions, and choroid plexus were normalized to the total intracranial volume.

^*^Significance level at *P*<0.05

*BMI* body mass index*, MCI* mild cognitive impairment, *MMSE* Mini-Mental State Examination, *SF* Sylvian fissure, *SHM* subarachnoid space at the high convexity and midline, *VS* ventricular system, *WMH* white matter hyperintensity.

**Table S6: Results of hierarchical multiple regression analysis for MMSE score with volumes of brain structures**

|  | **All participants (*n*=1,370)** | | | |  | **Participants without MCI (*n*=1,144)** | | | |
| --- | --- | --- | --- | --- | --- | --- | --- | --- | --- |
| Rankk | Brain structure | β_STD_ | *P*-value | ΔR^2^ | Rankk | Brain structure | β_STD_ | *P*-value | ΔR^2^ |
| 1 | Choroid plexus | −0.1097 | <0.0001* | 0.0096 | 1 | Choroid plexus | −0.1086 | 0.0006* | 0.0094 |
| 2 | SHM | 0.0930 | 0.0004* | 0.0078 | 2 | Transverse temporal | 0.0797 | 0.0092 | 0.0054 |
| 3 | VS | −0.0815 | 0.0033 | 0.0054 | 3 | Supramarginal | 0.0741 | 0.0199 | 0.0043 |
| 4 | Middle temporal | 0.0832 | 0.0040 | 0.0051 | 4 | Superior frontal | 0.0738 | 0.0290 | 0.0038 |
| 5 | Supramarginal | 0.0748 | 0.0085 | 0.0043 | 5 | SF | −0.0650 | 0.0346 | 0.0036 |
| 6 | Accumbens | 0.0753 | 0.0093 | 0.0042 | 6 | Accumbens | 0.0648 | 0.0473 | 0.0031 |
| 7 | Hippocampus | 0.0770 | 0.0110 | 0.0040 | 7 | VS | −0.0615 | 0.0481 | 0.0031 |
| 8 | Lateral occipital | 0.0717 | 0.0118 | 0.0039 | 8 | SHM | 0.0529 | 0.0746 | 0.0025 |
| 9 | SF | −0.0676 | 0.0130 | 0.0038 | 9 | Banks sts | 0.0569 | 0.0826 | 0.0024 |
| 10 | Amygdala | 0.0718 | 0.0131 | 0.0038 | 10 | Precentral | 0.0654 | 0.0834 | 0.0024 |
| 11 | Precuneus | 0.0745 | 0.0132 | 0.0038 | 11 | Lateral occipital | 0.0548 | 0.0866 | 0.0024 |
| 12 | Lingual | 0.0669 | 0.0153 | 0.0037 | 12 | Temporal pole | −0.0484 | 0.1082 | 0.0021 |
| 13 | Transverse temporal | 0.0641 | 0.0185 | 0.0034 | 13 | Thalamus | 0.0429 | 0.1814 | 0.0014 |
| 14 | Superior frontal | 0.0678 | 0.0260 | 0.0031 | 14 | Inferior parietal | 0.0433 | 0.1995 | 0.0013 |
| 15 | Lateral orbitofrontal | 0.0608 | 0.0300 | 0.0029 | 15 | Superior temporal | 0.0396 | 0.2256 | 0.0012 |
| 16 | Banks sts | 0.0611 | 0.0347 | 0.0028 | 16 | Lingual | 0.0350 | 0.2613 | 0.0010 |
| 17 | Superior temporal | 0.0612 | 0.0353 | 0.0028 | 17 | Inferior temporal | 0.0333 | 0.2905 | 0.0009 |
| 18 | Entorhinal | 0.0515 | 0.0507 | 0.0024 | 18 | Fusiform | 0.0328 | 0.2939 | 0.0009 |
| 19 | Inferior parietal | 0.0586 | 0.0515 | 0.0024 | 19 | Precuneus | 0.0330 | 0.3276 | 0.0008 |
| 20 | Inferior temporal | 0.0510 | 0.0691 | 0.0021 | 20 | Pars opercularis | 0.0290 | 0.3372 | 0.0007 |
| 21 | Precentral | 0.0576 | 0.0878 | 0.0018 | 21 | Entorhinal | −0.0272 | 0.3555 | 0.0007 |
| 22 | Fusiform | 0.0443 | 0.1126 | 0.0016 | 22 | Pars triangularis | 0.0274 | 0.3612 | 0.0007 |
| 23 | Insula | 0.0401 | 0.1671 | 0.0012 | 23 | Lateral orbitofrontal | 0.0268 | 0.3922 | 0.0006 |
| 24 | Thalamus | 0.0395 | 0.1677 | 0.0012 | 24 | Pallidum | 0.0246 | 0.4319 | 0.0005 |
| 25 | Pars triangularis | 0.0362 | 0.1750 | 0.0011 | 25 | Pars orbitalis | 0.0219 | 0.4717 | 0.0004 |
| 26 | Pars opercularis | 0.0332 | 0.2161 | 0.0010 | 26 | Postcentral | 0.0214 | 0.5065 | 0.0004 |
| 27 | Parahippocampal | 0.0338 | 0.2298 | 0.0009 | 27 | Paracentral | 0.0217 | 0.5088 | 0.0003 |
| 28 | Pallidum | 0.0316 | 0.2472 | 0.0008 | 28 | Middle temporal | 0.0208 | 0.5167 | 0.0003 |
| 29 | Pars orbitalis | 0.0311 | 0.2512 | 0.0008 | 29 | Pericalcarine | −0.0146 | 0.6183 | 0.0002 |
| 30 | Posterior cingulate | 0.0280 | 0.3181 | 0.0006 | 30 | Rostral middle frontal | 0.0129 | 0.6727 | 0.0001 |
| 31 | Postcentral | 0.0251 | 0.3834 | 0.0005 | 31 | Parahippocampal | 0.0120 | 0.7061 | 0.0001 |
| 32 | Caudate | 0.0234 | 0.4209 | 0.0004 | 32 | Caudal anterior cingulate | −0.0102 | 0.7325 | 0.0001 |
| 33 | Rostral middle frontal | 0.0191 | 0.4843 | 0.0003 | 33 | Isthmus cingulate | −0.0092 | 0.7625 | 0.0001 |
| 34 | Superior parietal | 0.0209 | 0.4923 | 0.0003 | 34 | Insula | 0.0095 | 0.7691 | 0.0001 |
| 35 | Putamen | 0.0174 | 0.5255 | 0.0003 | 35 | Superior parietal | −0.0096 | 0.7791 | 0.0001 |
| 36 | Rostral anterior cingulate | 0.0145 | 0.5798 | 0.0002 | 36 | Cuneus | −0.0073 | 0.8071 | 0.0000 |
| 37 | Caudal middle frontal | 0.0151 | 0.5880 | 0.0002 | 37 | Putamen | 0.0074 | 0.8122 | 0.0000 |
| 38 | Isthmus cingulate | 0.0142 | 0.5987 | 0.0002 | 38 | Medial orbitofrontal | −0.0064 | 0.8278 | 0.0000 |
| 39 | Cuneus | 0.0126 | 0.6322 | 0.0001 | 39 | Rostral anterior cingulate | −0.0057 | 0.8489 | 0.0000 |
| 40 | Temporal pole | −0.0080 | 0.7637 | 0.0001 | 40 | Amygdala | −0.0055 | 0.8657 | 0.0000 |
| 41 | Pericalcarine | 0.0078 | 0.7641 | 0.0001 | 41 | Frontal pole | 0.0044 | 0.8848 | 0.0000 |
| 42 | Paracentral | −0.0066 | 0.8173 | 0.0000 | 42 | Caudate | 0.0037 | 0.9099 | 0.0000 |
| 43 | Frontal pole | −0.0041 | 0.8804 | 0.0000 | 43 | Posterior cingulate | −0.0031 | 0.9215 | 0.0000 |
| 44 | Caudal anterior cingulate | −0.0029 | 0.9110 | 0.0000 | 44 | Hippocampus | 0.0025 | 0.9415 | 0.0000 |
| 45 | Medial orbitofrontal | 0.0003 | 0.9901 | 0.0000 | 45 | Caudal middle frontal | −0.0018 | 0.9550 | 0.0000 |

As the independent variables, each brain structure was separately added to the basic model, which included age, sex, education, hypertension, DM, dyslipidemia, atrial fibrillation, coronary artery disease, heart failure, BMI, history of smoking, MRI scanner, Fazekas score, lacunar infarction, microbleeds, and perivascular space.

For each analysis, the standardized regression coefficient (β_STD_) and change in R^2^ (∆R^2^) were calculated. Forty-five brain structures are shown in descending order of ∆R^2^.

*The significance level was P<0.0011 to correct for 45 modeling analyses.

BMI body mass index, MCI mild cognitive impairment, MMSE Mini-Mental State Examination, SF Sylvian fissure, SHM subarachnoid space at high convexity and midline, sts superior temporal sulcus, VS ventricular system.

**Table S7: Results of hierarchical multiple regression analysis for choroid plexus volume with volumes of other brain structures**

|  | **All participants (*n*=1,370)** | | | |  | **Participants without MCI (*n*=1,144)** | | | |
| --- | --- | --- | --- | --- | --- | --- | --- | --- | --- |
| Rank | Brain structure | β_STD_ | *P*-value | ΔR^2^ | Rank | Brain structure | β_STD_ | *P*-value | ΔR^2^ |
| 1 | VS | 0.4996 | <0.0001* | 0.2019 | 1 | VS | 0.5019 | <0.0001* | 0.2082 |
| 2 | SHM | −0.2997 | <0.0001* | 0.0812 | 2 | SHM | −0.2992 | <0.0001* | 0.0814 |
| 3 | Accumbens | −0.2490 | <0.0001* | 0.0459 | 3 | SF | 0.2463 | <0.0001* | 0.0513 |
| 4 | SF | 0.2202 | <0.0001* | 0.0406 | 4 | Accumbens | −0.2414 | <0.0001* | 0.0436 |
| 5 | Hippocampus | −0.2293 | <0.0001* | 0.0356 | 5 | Hippocampus | −0.2090 | <0.0001* | 0.0305 |
| 6 | Amygdala | −0.1888 | <0.0001* | 0.0264 | 6 | Amygdala | −0.1726 | <0.0001* | 0.0228 |
| 7 | Inferior temporal | −0.1827 | <0.0001* | 0.0263 | 7 | Posterior cingulate | −0.1553 | <0.0001* | 0.0196 |
| 8 | Posterior cingulate | −0.1803 | <0.0001* | 0.0257 | 8 | Inferior temporal | −0.1536 | <0.0001* | 0.0191 |
| 9 | Supramarginal | −0.1676 | <0.0001* | 0.0216 | 9 | Inferior parietal | −0.1510 | <0.0001* | 0.0160 |
| 10 | Lateral orbitofrontal | −0.1648 | <0.0001* | 0.0215 | 10 | Lateral orbitofrontal | −0.1395 | <0.0001* | 0.0159 |
| 11 | Superior temporal | −0.1668 | <0.0001* | 0.0204 | 11 | Lateral occipital | −0.1383 | <0.0001* | 0.0150 |
| 12 | Fusiform | −0.1518 | <0.0001* | 0.0184 | 12 | Parahippocampal | −0.1346 | <0.0001* | 0.0144 |
| 13 | Inferior parietal | −0.1614 | <0.0001* | 0.0179 | 13 | Supramarginal | −0.1349 | <0.0001* | 0.0144 |
| 14 | Lateral occipital | −0.1450 | <0.0001* | 0.0161 | 14 | Superior temporal | −0.1377 | <0.0001* | 0.0142 |
| 15 | Parahippocampal | −0.1410 | <0.0001* | 0.0156 | 15 | Rostral middle frontal | −0.1270 | <0.0001* | 0.0139 |
| 16 | Superior frontal | −0.1506 | <0.0001* | 0.0152 | 16 | Superior frontal | −0.1352 | <0.0001* | 0.0128 |
| 17 | Precuneus | −0.1448 | <0.0001* | 0.0144 | 17 | Fusiform | −0.1099 | 0.0002* | 0.0099 |
| 18 | Rostral middle frontal | −0.1243 | <0.0001* | 0.0129 | 18 | Precuneus | −0.1167 | 0.0002* | 0.0096 |
| 19 | Pars orbitalis | −0.1216 | <0.0001* | 0.0125 | 19 | Caudate | 0.1129 | 0.0003* | 0.0094 |
| 20 | Caudate | 0.1306 | <0.0001* | 0.0125 | 20 | Pars orbitalis | −0.1045 | 0.0003* | 0.0094 |
| 21 | Medial orbitofrontal | −0.1154 | <0.0001* | 0.0121 | 21 | Medial orbitofrontal | −0.1010 | 0.0003* | 0.0093 |
| 22 | Pars triangularis | −0.1125 | <0.0001* | 0.0111 | 22 | Transverse temporal | −0.1003 | 0.0005* | 0.0086 |
| 23 | Postcentral | −0.1210 | <0.0001* | 0.0110 | 23 | Pars triangularis | −0.0959 | 0.0007* | 0.0082 |
| 24 | Putamen | −0.1127 | <0.0001* | 0.0105 | 24 | Putamen | −0.0976 | 0.0009* | 0.0078 |
| 25 | Caudal anterior cingulate | −0.1074 | <0.0001* | 0.0104 | 25 | Caudal anterior cingulate | −0.0903 | 0.0013 | 0.0073 |
| 26 | Transverse temporal | −0.1097 | <0.0001* | 0.0101 | 26 | Isthmus cingulate | −0.0908 | 0.0016 | 0.0071 |
| 27 | Middle temporal | −0.1090 | 0.0001* | 0.0088 | 27 | Postcentral | −0.0942 | 0.0019 | 0.0068 |
| 28 | Temporal pole | −0.0953 | 0.0002* | 0.0080 | 28 | Lingual | −0.0901 | 0.0021 | 0.0067 |
| 29 | Isthmus cingulate | −0.0970 | 0.0002* | 0.0080 | 29 | Precentral | −0.1054 | 0.0030 | 0.0062 |
| 30 | Insula | −0.1031 | 0.0003* | 0.0079 | 30 | Pallidum | −0.0835 | 0.0045 | 0.0057 |
| 31 | Superior parietal | −0.1030 | 0.0005* | 0.0079 | 31 | Banks sts | −0.0861 | 0.0052 | 0.0055 |
| 32 | Precentral | −0.1109 | 0.0007* | 0.0071 | 32 | Middle temporal | −0.0808 | 0.0073 | 0.0051 |
| 33 | Thalamus | −0.0892 | 0.0013 | 0.0067 | 33 | Thalamus | −0.0809 | 0.0074 | 0.0051 |
| 34 | Banks sts | −0.0899 | 0.0014 | 0.0060 | 34 | Rostral anterior cingulate | −0.0748 | 0.0075 | 0.0051 |
| 35 | Rostral anterior cingulate | −0.0804 | 0.0016 | 0.0060 | 35 | Temporal pole | −0.0751 | 0.0081 | 0.0050 |
| 36 | Pallidum | −0.0836 | 0.0016 | 0.0058 | 36 | Insula | −0.0759 | 0.0129 | 0.0044 |
| 37 | Lingual | −0.0828 | 0.0020 | 0.0058 | 37 | Superior parietal | −0.0752 | 0.0199 | 0.0038 |
| 38 | Caudal middle frontal | −0.0739 | 0.0063 | 0.0056 | 38 | Caudal middle frontal | −0.0624 | 0.0337 | 0.0032 |
| 39 | Frontal pole | −0.0701 | 0.0075 | 0.0044 | 39 | Frontal pole | −0.0522 | 0.0708 | 0.0023 |
| 40 | Pars opercularis | −0.0576 | 0.0276 | 0.0042 | 40 | Pars opercularis | −0.0511 | 0.0728 | 0.0023 |
| 41 | Entorhinal | −0.0548 | 0.0328 | 0.0029 | 41 | Pericalcarine | 0.0397 | 0.1484 | 0.0015 |
| 42 | Pericalcarine | 0.0369 | 0.1435 | 0.0027 | 42 | Paracentral | −0.0373 | 0.2267 | 0.0010 |
| 43 | Paracentral | −0.0314 | 0.2619 | 0.0013 | 43 | Entorhinal | −0.0178 | 0.5208 | 0.0003 |
| 44 | Cuneus | −0.0219 | 0.3925 | 0.0004 | 44 | Cuneus | −0.0124 | 0.6589 | 0.0001 |

As the independent variables, each brain structure was separately added to the basic model, which included age, sex, education, hypertension, DM, dyslipidemia, atrial fibrillation, coronary artery disease, heart failure, BMI, history of smoking, MRI scanner, Fazekas score, lacunar infarction, microbleeds, and perivascular space.

For each analysis, the standardized regression coefficient (β_STD_) and change in R^2^ (∆R^2^) were calculated. Forty-four brain structures are shown in descending order of ∆R^2^.

*The significance level was P<0.0011 to correct for 44 modeling analyses.

BMI body mass index, MCI mild cognitive impairment, MMSE Mini-Mental State Examination, SF Sylvian fissure, SHM subarachnoid space at high convexity and midline, sts superior temporal sulcus, VS ventricular system.
